# Increased risk of SARS-CoV-2 reinfection associated with emergence of Omicron in South Africa

**DOI:** 10.1101/2021.11.11.21266068

**Authors:** Juliet R.C. Pulliam, Cari van Schalkwyk, Nevashan Govender, Anne von Gottberg, Cheryl Cohen, Michelle J. Groome, Jonathan Dushoff, Koleka Mlisana, Harry Moultrie

## Abstract

**Introduction:** Globally, there have been more than 404 million cases of SARS-CoV-2, with 5.8 million confirmed deaths, as of February 2022. South Africa has experienced four waves of SARS-CoV-2 transmission, with the second, third, and fourth waves being driven by the Beta, Delta, and Omicron variants, respectively. A key question with the emergence of new variants is the extent to which they are able to reinfect those who have had a prior natural infection.

**Rationale:** We developed two approaches to monitor routine epidemiological surveillance data to examine whether SARS-CoV-2 reinfection risk has changed through time in South Africa, in the context of the emergence of the Beta (B.1.351), Delta (B.1.617.2), and Omicron (B.1.1.529) variants. We analyze line list data on positive tests for SARS-CoV-2 with specimen receipt dates between 04 March 2020 and 31 January 2022, collected through South Africa’s National Notifiable Medical Conditions Surveillance System. Individuals having sequential positive tests at least 90 days apart were considered to have suspected reinfections. Our routine monitoring of reinfection risk included comparison of reinfection rates to the expectation under a null model (approach 1) and estimation of the time-varying hazards of infection and reinfection throughout the epidemic (approach 2) based on model-based reconstruction of the susceptible populations eligible for primary and second infections.

**Results:** 105,323 suspected reinfections were identified among 2,942,248 individuals with laboratory-confirmed SARS-CoV-2 who had a positive test result at least 90 days prior to 31 January 2022. The number of reinfections observed through the end of the third wave in September 2021 was consistent with the null model of no change in reinfection risk (approach 1). Although increases in the hazard of primary infection were observed following the introduction of both the Beta and Delta variants, no corresponding increase was observed in the reinfection hazard (approach 2). Contrary to expectation, the estimated hazard ratio for reinfection versus primary infection was lower during waves driven by the Beta and Delta variants than for the first wave (relative hazard ratio for wave 2 versus wave 1: 0.71 (CI_95_: 0.60–0.85); for wave 3 versus wave 1: 0.54 (CI_95_: 0.45–0.64)). In contrast, the recent spread of the Omicron variant has been associated with an increase in reinfection hazard coefficient. The estimated hazard ratio for reinfection versus primary infection versus wave 1 was 1.75 (CI_95_: 1.48–2.10) for the period of Omicron emergence (01 November 2021 to 30 November 2021) and 1.70 (CI_95_: 1.44–2.04) for wave 4 versus wave 1. Individuals with identified reinfections since 01 November 2021 had experienced primary infections in all three prior waves, and an increase in third infections has been detected since mid-November 2021. Many individuals experiencing third infections had second infections during the third (Delta) wave that ended in September 2021, strongly suggesting that these infections resulted from immune evasion rather than waning immunity.

**Conclusion:** Population-level evidence suggests that the Omicron variant is associated with substantial ability to evade immunity from prior infection. In contrast, there is no population-wide epidemiological evidence of immune escape associated with the Beta or Delta variants. This finding has important implications for public health planning, particularly in countries like South Africa with high rates of immunity from prior infection. Further development of methods to track reinfection risk during pathogen emergence, including refinements to assess the impact of waning immunity, account for vaccine-derived protection, and monitor the risk of multiple reinfections will be an important tool for future pandemic preparedness.

## Introduction

As of 31 January 2022, South Africa had more than 3.6 million cumulative laboratory-confirmed cases of SARS-CoV-2, concentrated in four waves of infection (Figure 1). The first case was detected in early March 2020 and was followed by a wave that peaked in July 2020 and ended in September. The second wave, which peaked in January 2021 and ended in February, was driven by the Beta (B.1.351 / 501Y.V2 / 20H) variant, which was first detected in South Africa in October 2020 (*1*). The third wave, which peaked in July and ended in September 2021, was dominated by the Delta (B.1.617.2 / 478K.V1 / 21A) variant (*2*). In late November 2021, the Omicron (B.1.1.529 / 21K) variant was detected in Gauteng Province, the smallest yet most populous province, and associated with rapidly increasing case numbers (*3*). The estimated effective reproduction number in Gauteng based on PCR-confirmed cases was 2.3 as of 18 November, which was as high as had been seen at any point during prior three waves, and peaked above 3 in late November (*4, 5*). The proportion of positive PCR tests with S-gene target failure (SGTF), a marker of the BA.1 sublineage of the Omicron variant, subsequently increased across all provinces (*6*).

**Figure 1.**
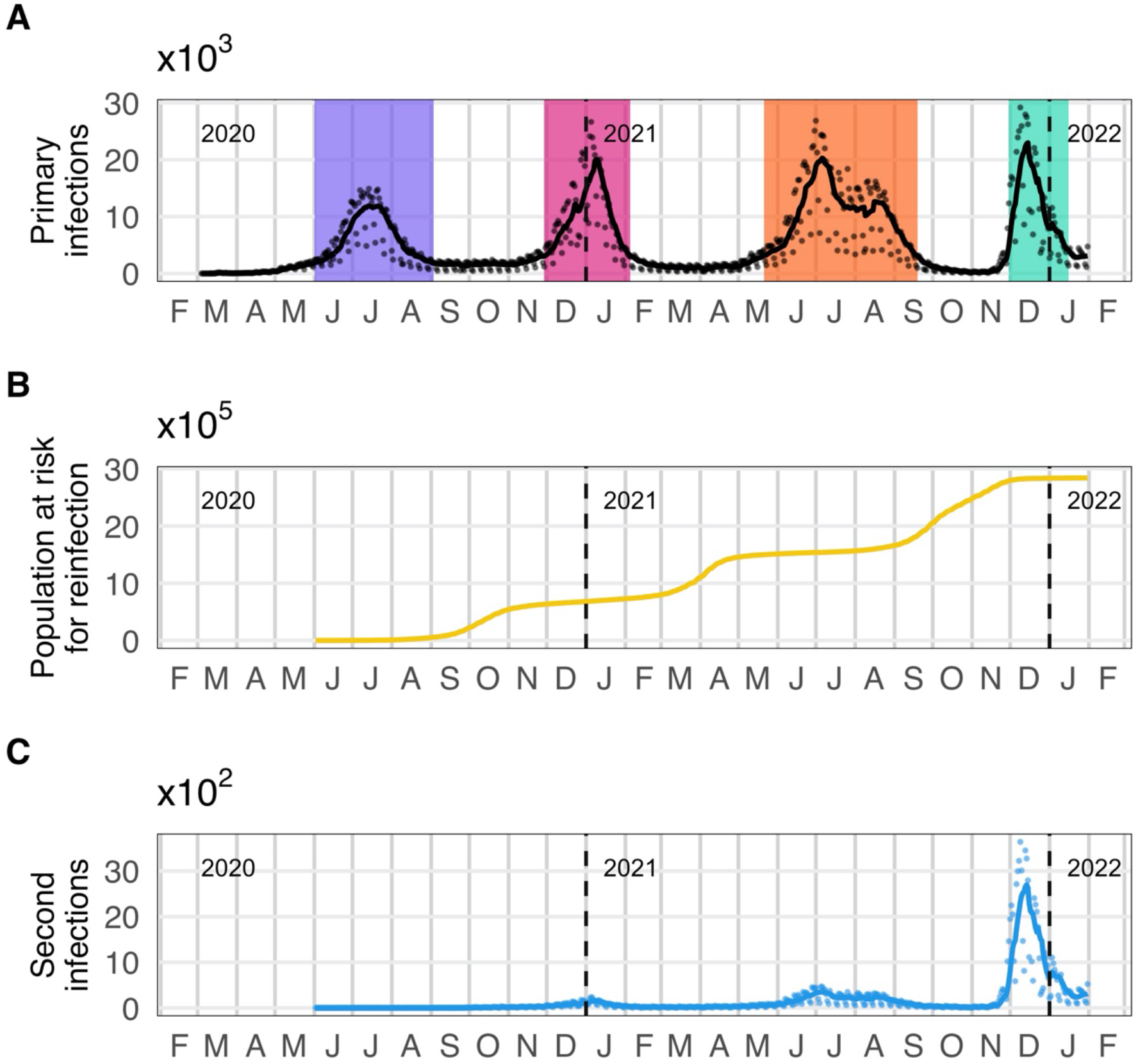
Daily numbers of detected primary infections, individuals eligible to be considered for reinfection, and suspected reinfections in South Africa. A: Time series of detected primary infections. Black line indicates 7-day moving average; black points are daily values. Colored bands represent wave periods, defined as the period for which the 7-day moving average of cases was at least 15% of the corresponding wave peak (purple = wave 1, pink = wave 2, orange = wave 3, turquoise = wave 4). B: Population at risk for reinfection (individuals whose most recent positive test was at least 90 days ago and who have not yet had a suspected reinfection). C: Time series of suspected reinfections. Blue line indicates 7-day moving average; blue points are daily values.

Following emergence of three variants of concern in South Africa, a key question remains of whether there is epidemiologic evidence of increased risk of SARS-CoV-2 reinfection with these variants (i.e., immune escape from natural infection). Laboratory-based studies suggest that convalescent serum has a reduced neutralizing effect on the Beta, Delta, and Omicron variants compared to wild type virus *in vitro* (*7–12*); however, this finding does not necessarily translate into immune evasion at the population level.

To examine whether reinfection risk has changed through time, it is essential to account for potential confounding factors affecting the incidence of reinfection: namely, the changing force of infection experienced by all individuals in the population and the growing number of individuals eligible for reinfection through time. These factors are tightly linked to the timing of epidemic waves. We examine reinfection trends in South Africa using two approaches that account for these factors to address the question of whether circulation of variants of concern has been associated with increased reinfection risk, as would be expected if their emergence was driven or facilitated by immune evasion.

## Identification of and characterization of reinfections

We define a suspected reinfection as a positive SARS-CoV-2 test in an individual with at least one previous positive test and whose most recent positive test occurred at least 90 days earlier. Based on routinely collected line list data maintained by the National Institute for Communicable Diseases (NICD) with specimen receipt dates between 04 March 2020 and 31 January 2022, we identified 105,323 individuals with at least two suspected infections, 1,778 individuals with at least three suspected infections, and 18 individuals with four suspected infections.

### Time between successive positive tests

The distribution of times between successive positive tests for individuals’ first and second infections has peaks near 170, 350, and 520 days (Figure 2A). The shape of the distribution was strongly influenced by the timing of South Africa’s epidemic waves, which have been spaced roughly six months apart. The first peak corresponds mainly to individuals whose primary infection and second infection occurred in consecutive waves (e.g., initially infected in wave 1 and reinfected in wave 2, initially infected in wave 2 and reinfected in wave 3, or initially infected in wave 3 and reinfected in wave 4), while the second peak corresponds mainly to individuals initially infected in wave 1 and reinfected in wave 3 or initially infected in wave 2 and reinfected in wave 4. The third peak corresponds to individuals initially infected in wave 1 and reinfected in wave 4.

**Figure 2.**
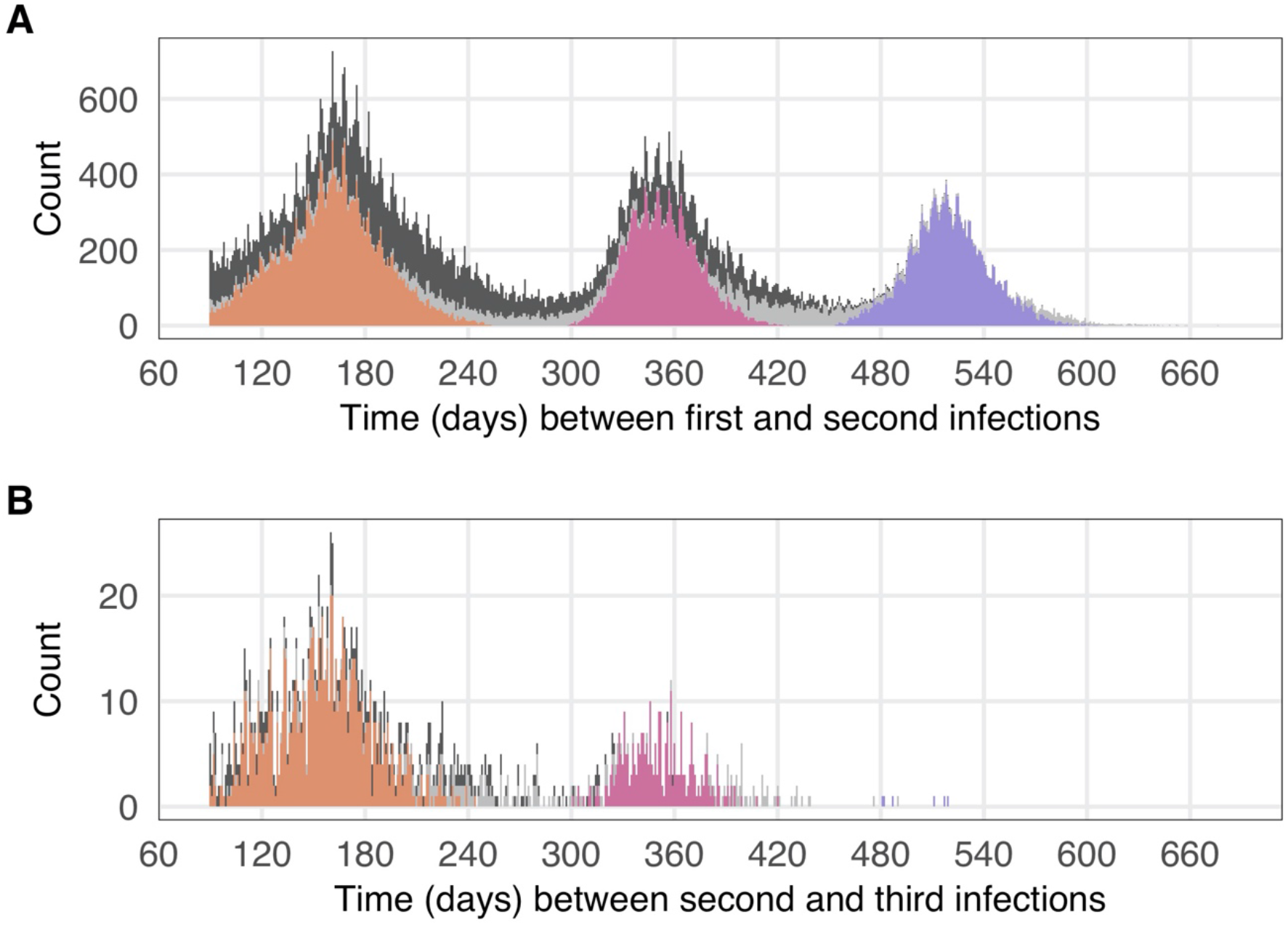
Time between consecutive infections, based on the time between successive positive tests. Note that the time since the previous positive test must be at least 90 days to be considered a reinfection. A: Time in days between the last positive test of the first infection and the first positive tests of the suspected second infection. B: Time in days between the last positive test of the putative second infection and the first positive tests of the suspected third infection. Colors represent suspected reinfections diagnosed on or after 01 November 2021. In both panels, bars for these individuals are colored by the wave during which the previous infection occurred (purple = wave 1, pink = wave 2, orange = wave 3, light grey = inter-wave).

Almost all suspected third infections occurred after 31 October 2021, i.e., during the period of Omicron circulation. The distribution of times between successive positive tests for individuals’ second and third infections has peaks corresponding to those whose second infections occurred in the second and third waves.

### Individuals with multiple suspected reinfections

1,778 individuals were identified who had three or more suspected infections. Prior to the emergence of Omicron, most of these individuals initially tested positive during the first wave, with suspected reinfections associated with waves two and three; however, 1,492 individuals with multiple reinfections (83.9%) experienced their third infection after 31 October 2021, which suggests that most third infections are associated with transmission of the Omicron variant (Figure 3).

**Figure 3.**
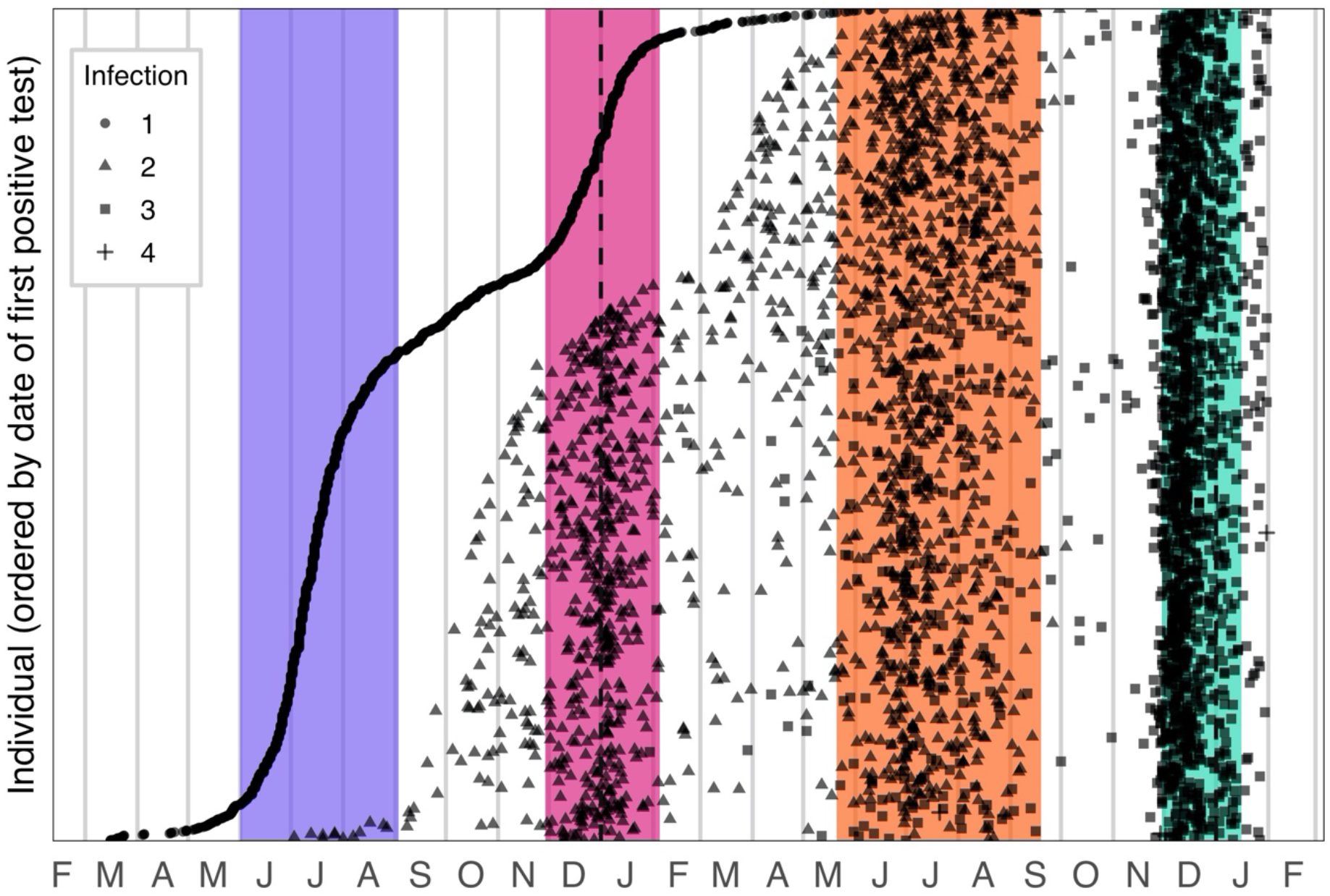
Timing of infections for individuals with multiple suspected reinfections. Circles represent the first positive test of the first detected infection; triangles represent the first positive test of the suspected second infection; squares represent the first positive test of the suspected third infection; crosses represent the first positive test of the suspected fourth infection. Colored bands represent wave periods, defined as the period for which the 7-day moving average of cases was at least 15% of the corresponding wave peak (purple = wave 1, pink = wave 2, orange = wave 3, turquoise = wave 4).

## Population-level reinfection trends in South Africa

The population at risk of reinfection has risen monotonically since the beginning of the epidemic, with relatively rapid increases associated with each wave (delayed by 90 days because of our definition of reinfection, Figure 1B). No suspected reinfections were detected until 23 June 2020, after which the number of suspected reinfections increased gradually. The 7-day moving average of suspected second infections reached a peak of approximately 160 during the second epidemic wave and 350 during the third wave (Figure 1). Following the third wave, the number of reinfections began to increase dramatically in mid-November 2022. During the fourth wave, the 7-day moving average of suspected second infections reached nearly 2,700 and the 7-day moving average of all suspected reinfections (including second, third, and fourth infections) reached approximately 2,750.

### Comparison of data to projections from a null model

We developed a catalytic model to project the expected number of reinfections through time under the assumption of a constant reinfection hazard coefficient (i.e., a null model of no change in reinfection risk). The model assumes the reinfection hazard is proportional to the 7-day moving average of the total number of diagnosed infections (primary infections and reinfections). During our early monitoring of reinfection risk, we fitted the reinfection hazard coefficient to data from 02 June 2020 to 30 September 2020 to parameterize the null model of no change in the reinfection hazard coefficient through time, and projected the number of reinfections through 30 June 2021. Based on this, we concluded there was no population-level evidence of immune escape and recommended on-going monitoring of reinfection trends (*13*).

Given that there was no evidence of divergence from the null projection during the second wave, and to improve convergence of the MCMC fitting algorithm, for the present analysis, we repeated the fitting process using a window of 02 June 2020 to 28 February 2021 (representing the end of the month in which the second wave ended). This led to good convergence with regard to estimation of both the negative binomial dispersion parameter and the reinfection hazard coefficient (Figure S4) and allowed us to fit the model to all nine provinces. The 7-day moving average of observed reinfections and most individual daily values fall within the projection interval from the beginning of the projection period though the end of the third wave (Figure 4). From early November 2021, however, the 7-day moving average of observed reinfections reached the upper bound of the projection interval, with many individual daily numbers falling well above the projection interval, both nationally and in Gauteng (Figure 4). This observed deviation from the projection under the null model is a signature of immune evasion, and the timing of this deviation suggests it is associated with the emergence of the Omicron variant. A similar pattern has now been seen across all provinces (Figures S5-S7).

**Figure 4.**
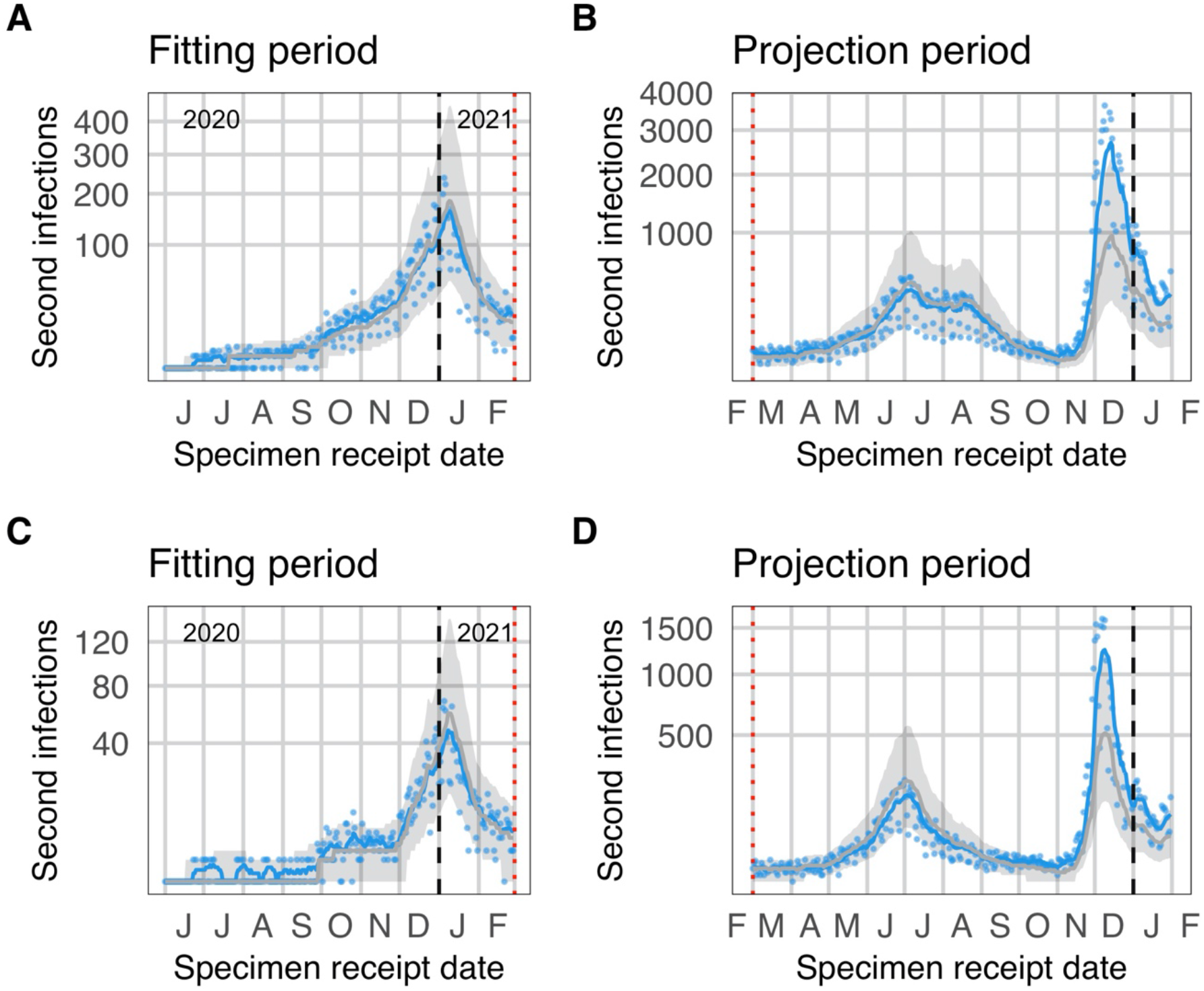
Observed and expected temporal trends in reinfection numbers. Blue lines (points) represent the 7-day moving average (daily values) of suspected reinfections. Grey lines (bands) represent mean predictions (95% projection intervals) from the null model. The null model was fit to data on suspected reinfections through 28 February 2021. Comparison of data to projections from the null model over the projection period. The divergence observed reinfections from the projection interval in November is suggestive of immune escape. A and B: National. C and D: Gauteng.

### Estimation of time-varying infection and reinfection hazards

We also examined changes in the reinfection risk via a method that relies on reconstruction of the numbers of observed and unobserved first and second infections through time (see Materials and Methods for details). Based on this approach, the estimated hazard coefficient for primary infection increased steadily through the end of the third wave, as expected under a combination of relaxing of restrictions, behavioral fatigue, and introduction of variants with increased transmissibility (Beta and Delta). The estimated hazard coefficient for reinfection, in contrast, remained relatively constant throughout this period, with the exception of an initial spike in mid-2020 (Figure 5). Because both reinfection numbers and the population eligible for reinfection were very low at the time, this increase may be an artifact of intense follow-up of the earliest cases or simply noise due to small numbers. The mean ratio of reinfection hazard to primary infection hazard decreased slightly from 0.15 in wave 1 to 0.12 in wave 2 and 0.09 in wave 3. The absolute values of the hazard coefficients and hazard ratio are sensitive to assumed observation probabilities for primary infections and reinfections; however, the temporal trends are robust (Figure S8).

**Figure 5.**
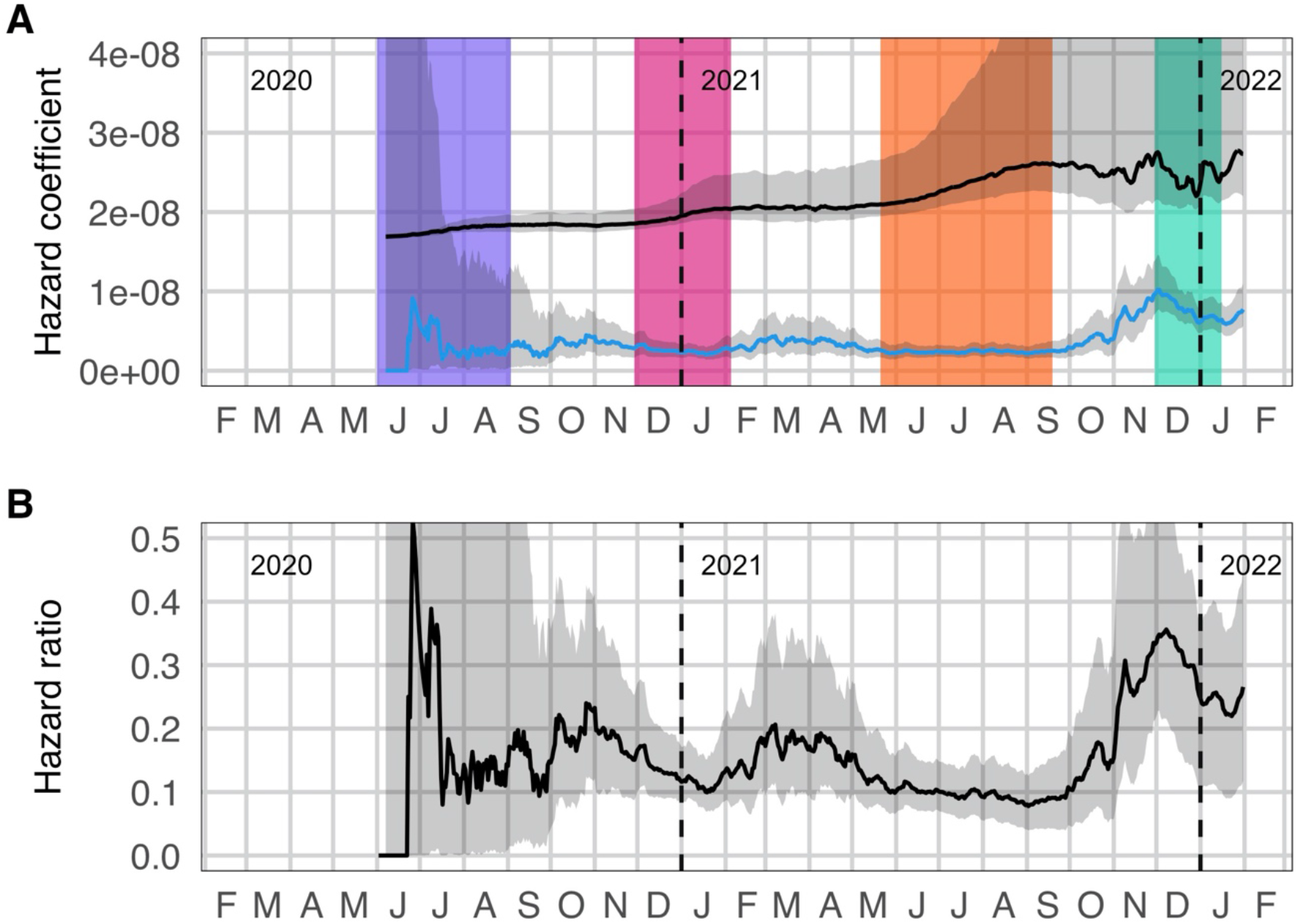
Estimates of infection and reinfection hazards. A: Estimated time-varying hazard coefficients for primary infection (black) and second infections (blue). Colored bands represent wave periods, defined as the period for which the 7-day moving average of cases was at least 15% of the corresponding wave peak (purple = wave 1, pink = wave 2, orange = wave 3, turquoise = wave 4). B: Ratio of the empirical hazard for reinfections to the empirical hazard for primary infections.

The picture changed following the end of the third wave. Although there is substantial uncertainty in the estimated hazard coefficient for primary infection, it appeared to decrease from early October 2021, with a simultaneous increase in the estimated reinfection hazard coefficient (Figure 5). This change became more marked from the beginning of November, with the mean ratio of reinfection hazard to primary infection hazard for the period from 01 November 2021 to the beginning of the fourth wave increasing to 0.25, and a mean ratio during the fourth wave of 0.27.

These findings are consistent with the estimates from the generalized linear mixed model based on the reconstructed data set. In this analysis, the *relative* hazard ratio for wave 2 versus wave 1 was 0.71 (CI_95_: 0.60–0.85) and for wave 3 versus wave 1 was 0.54 (CI_95_: 0.45–0.64). The relative hazard ratio for the period of Omicron emergence (01 November 2021 to the start of the fourth wave) versus wave 1 was 1.75 (CI_95_: 1.48–2.10), and for wave 4 versus wave 1 was 1.70 (CI_95_: 1.44–2.04).

## Discussion and limitations

Our analyses suggest that the cumulative number of reinfections observed through the end of wave 3 was consistent with the null model of no change in reinfection risk through time. Furthermore, our findings suggest that the relative hazard of reinfection versus primary infection decreased with each subsequent wave of infections through September 2021, as would be expected if the risk of primary infection increased without a corresponding increase in reinfection risk. Thus, our analyses show no population-level evidence of immune escape associated with emergence of the Beta or Delta variants. In contrast, in November 2021, the number of daily new reinfections spiked and exceeded the 95% projection interval from the null model, accompanied by a notable increase in the hazard ratio for reinfection versus primary infection. The timing of these changes strongly suggests that they were driven by the emergence of the Omicron variant.

Differences in the time-varying force of infection, original and subsequent circulating lineages, testing strategies, and vaccine coverage limit the usefulness of direct comparisons of rates of reinfections across countries or studies. Pre-Omicron reinfection does however appear to be relatively uncommon. The PCR-confirmed reinfection rate ranged from 0% – 1.1% across eleven studies included in a systematic review (*14*). While none of the studies included in the systematic review reported increasing risk of reinfection over time, the duration of follow-up was less than a year and most studies were completed prior to the identification of variants of concern, and all studies predated the emergence of Omicron. Our findings for the period prior to the emergence of Omicron are consistent with results from the PHIRST-C community cohort study conducted in two locations in South Africa, which found that infection prior to the second wave provided 84% protection against reinfection during the second (Beta) wave (*15*), comparable to estimates of the level of protection against reinfection for wild type virus from the SIREN study in the UK (*16*).

A preliminary analysis of reinfection trends in England suggested that the Delta variant may have a higher risk of reinfection compared to the Alpha variant (*17*); however, this analysis did not take into account the temporal trend in the population at risk for reinfection, which may have biased the findings.

Our findings regarding the Beta and Delta variants are somewhat at odds with *in vitro* neutralization studies. Both the Beta and Delta variants are associated with decreased neutralization by some anti-receptor binding-domain (anti-RBD) and anti-N-terminal domain (anti-NTD) monoclonal antibodies though both Beta and Delta each remain responsive to at least one anti-RBD (*8, 9, 18*). In addition, Beta and Delta are relatively poorly neutralized by convalescent sera obtained from unvaccinated individuals infected with non-VOC virus (*7–9, 18*). Lastly sera obtained from individuals after both one and two doses of the BNT162b2 (Pfizer) or ChAdOx1 (AstraZeneca) vaccines displayed lower neutralization of the Beta and Delta variants when compared to non-VOC and Alpha variant (*9*); although this does not have direct bearing on reinfection risk, it is an important consideration for evaluating immune escape more broadly. Non-neutralizing antibodies and T-cell responses could explain the apparent disjuncture between our findings and the *in vitro* immune evasion demonstrated by both Beta and Delta.

### Strengths of this study

Our study has three major strengths. First, we analyzed a large routine national data set comprising all confirmed cases in the country, allowing a comprehensive analysis of suspected reinfections in the country. Second, we found consistent results using two different analytical methods, both of which accounted for the changing force of infection and increasing numbers of individuals at risk for reinfection. Third, our real-time routine monitoring was sufficient to detect a population-level signal of immune evasion during the initial period of emergence of the Omicron variant in South Africa, prior to results from laboratory-based neutralization tests, providing timely information of importance to global public health planning.

### Limitations of this study

The primary limitation of this study is that changes in testing practices, health-seeking behavior, or access to care have not been accounted for in these analyses. Estimates based on serological data from blood donors suggest substantial geographic variability in detection rates (*19*), which may contribute to the observed differences in reinfection patterns by province (Supplementary Figure S1). Detection rates likely also vary through time and by other factors affecting access to testing, which may include occupation, age, and socioeconomic status. In particular, rapid antigen tests, which were introduced in South Africa in late 2020, may be under-reported despite mandatory reporting requirements. Although we have incorporated adjustments that account for late reporting of antigen tests, if under-reporting of antigen tests was substantial and time-varying it could still influence our findings. However, comparing temporal trends in infection risk among those eligible for reinfection with the rest of the population, as in approach 2, mitigates against potential failure to detect a substantial increase in risk.

Civil unrest during July 2021 severely disrupted testing in Gauteng and KwaZulu-Natal, the two most populous provinces in the country. Case data are unreliable during the period of unrest and a key assumption of our models - that the force of infection is proportional to the number of positive tests - was violated during this period, resulting in increased misclassification of individuals regarding their status as to whether they are at risk of primary or re-infection. The effect of this misclassification on the signal of immune escape during the period of Omicron’s emergence would likely be small and would be expected to bias subsequent reinfection hazard estimates downwards.

The purpose of our analysis is to detect changes in the relative reinfection risk through time, rather than to precisely estimate what the reinfection risk is at any particular point in time. While issues related to underdetection of both primary infections and reinfections are likely to affect the projection intervals against which we compare observed reinfections, we believe that our assessment of changes in the reinfection hazard are fairly robust to these detection issues. In effect, Approach 1 follows an open cohort of individuals who have had a first detected infection. Through time, this may include an increasing number of individuals whose first true infection was missed and whose first diagnosed infection is in fact a reinfection. These individuals would presumably be at a reduced risk of acquiring a new infection relative to those whose first detected infection was their first true infection. Two other factors would bias the results in the same direction: undetected reinfections in the cohort of individuals having had a first detected infection and deaths within this cohort, which are not accounted for due to not having a mortality line list that can be linked to the positive test data. All three factors artificially inflate the estimated denominator of individuals at risk for a second detected infection, thereby reducing the apparent reinfection risk. These factors may explain the slightly lower observed than projected number of reinfections throughout the Delta wave but did not have a substantial enough effect to prevent detection of the increased reinfection risk associated with the Omicron variant.

The other main limitation of this study is that reinfections were not confirmed by sequencing or by requiring a negative test between putative infections. Nevertheless, the 90-day window period between consecutive positive tests reduces the possibility that suspected reinfections were predominantly the result of prolonged viral shedding. Furthermore, due to data limitations, we were unable to examine whether symptoms and severity in primary episodes correlate with protection against subsequent reinfection.

Lastly, while vaccination may increase protection in previously infected individuals (*20–23*), vaccination coverage in South Africa was very low during much of the study period, with 22.5% of the population fully vaccinated by 30 November 2021 (*24*). Nevertheless, increasing vaccination uptake may reduce the risks of both primary infection and reinfection. The vaccination status of individuals with suspected reinfections identified in this study was unknown. Application of our approach to other locations with higher vaccine coverage would require a more nuanced consideration of the potential effect of vaccination. Further areas for future methodological development include accounting for potential of waning of natural and vaccine-derived immunity, as well as methods to track changes in the risk of multiple (three or more) infections.

Given the limitations outlined above, estimates of the extent of immune evasion based on our approach, which aims to detect changing trends rather than make precise estimates, should be treated with caution.

### Conclusion

We find evidence of a substantial increase in the risk of reinfection that is temporally consistent with the timing of the emergence of the Omicron variant in South Africa, suggesting that Omicron’s selection advantage is at least partially driven by an increased ability to infect previously infected individuals.

In contrast, we find no evidence that reinfection risk increased as a result of the emergence of Beta or Delta variants, suggesting that the selective advantage that allowed these variants to spread derived primarily from increased transmissibility, rather than immune evasion. The discrepancy between the population-level evidence presented here and expectations based on laboratory-based neutralization assays for Beta and Delta highlights the need to identify better correlates of immunity for assessing immune escape *in vitro*.

Immune evasion from prior infection has important implications for public health globally. As new variants emerge, methods to quantify the extent of immune evasion for both natural and vaccine-derived immunity, as well as changes in transmissibility and disease severity will be urgent priorities to inform facility readiness planning and other public health operations.

## Methods

### Data sources

Data analyzed in this study came from two sources maintained by the National Institute for Communicable Diseases (NICD): the outbreak response component of the Notifiable Medical Conditions Surveillance System (NMC-SS) deduplicated case list and the line list of repeated SARS-CoV-2 tests. All positive tests conducted in South Africa appear in the combined data set, regardless of the reason for testing or type of test (PCR or antigen detection), and include the large number of positive tests that were retrospectively added to the data set on 23 November 2021 (*25*). We note that, of the 18,585 cases reported on 23 November, 93% had a specimen receipt date before 01 November 2021, and 6% had specimen receipt dates on or after 21 November 2021.

A combination of deterministic (national identity number, names, dates of birth) and probabilistic linkage methods were utilized to identify repeated tests conducted on the same person. In addition, provincial COVID-19 contact tracing teams identify and report repeated SARS-CoV-2 positive tests to the NICD, whether detected via PCR or antigen tests. The unique COVID-19 case identifier which links all tests from the same person was used to merge the two datasets. Irreversibly hashed case IDs were generated for each individual in the merged data set.

Primary infections and suspected repeat infections were identified using the merged data set. Repeated case IDs in the line list were identified and used to calculate the time between consecutive positive tests for each individual, using specimen receipt dates. If the time between sequential positive tests was at least 90 days, the more recent positive test was considered to indicate a suspected new infection. We present a descriptive analysis of suspected third and fourth infections, although only suspected second infections (which we refer to as “reinfections”) were considered in the analyses of temporal trends. Incidence time series for primary infections and reinfections are calculated by specimen receipt date of the first positive test associated with the infection, and total observed incidence is calculated as the sum of first infections and reinfections. The specimen receipt date was chosen as the reference point for analysis because it is complete within the data set; however, problems have been identified with accuracy of specimen receipt dates for tests associated with substantially delayed reporting from some laboratories. For these tests, which had equivalent entries for specimen receipt date and specimen report date that were more than 7 days after the sample collection date, the specimen receipt date was adjusted to be 1 day after the sample collection date, reflecting the median delay across all tests.

All analyses were conducted in the R statistical programming language (R version 4.0.5 (2021-03-31)).

### Timing of reinfections

We calculated the time between successive infections as the number of days between the last positive test associated with an individual’s first identified infection (i.e., within 90 days of a previous positive test, if any) and the first positive test associated with their suspected subsequent infection (i.e., at least 90 days after the most recent positive test). We analyzed the distribution of these times for all second and third infections, and for the subset of second and third infections occurring since 01 November 2021.

### Statistical analysis of reinfection trends

We analysed the NICD national SARS-CoV-2 routine surveillance data to evaluate whether reinfection risk has changed since emergence of variants of concern in South Africa. We evaluated the daily numbers of suspected reinfections using two approaches. First, we constructed a simple null model based on the assumption that the reinfection hazard experienced by previously diagnosed individuals is proportional to the incidence of detected infections and fit this model to the pattern of reinfections observed through 28 February 2021. The null model assumes no change in the reinfection hazard coefficient through time. We then compared observed reinfections after the fitting period to expected reinfections under projections from the null model.

Second, we evaluated whether there has been a change in the *relative* hazard of reinfection versus primary infection, to distinguish between increased overall transmissibility of the variants and any *additional* risk of reinfection due to potential immune escape. To do this, we calculated a hazard coefficient at each time point for primary and second infections and compared their relative values through time.

#### Approach 1: Catalytic model assuming a constant reinfection hazard coefficient Model description

For a case testing positive on day *t* (by specimen receipt date), we assumed the reinfection hazard is 0 for each day from *t* + 1 to *t* + 89 and *λÎ*_*τ*_ for each day *τ* ≥ *t* + 90, where *Î*_*τ*_ is the 7-day moving average of the detected case incidence (first infections and reinfections) for day *τ*. The probability of a case testing positive on day *t* having a diagnosed reinfection by day *x* is thus 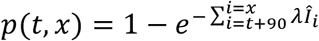, and the expected number of cases testing positive on day *t* that have had a diagnosed reinfection by day *x* is 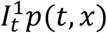, where 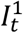 is the detected case incidence (putative first infections only) for day *t*. Thus, the expected cumulative number of reinfections by day *x* is 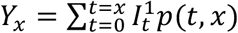. The expected daily incidence of reinfections on day *x* is *D*_*x*_ = *Y*_*x*_ – *Y*_*x*−1_.

#### Model fitting

The model was fitted to observed reinfection incidence through 28 February 2021 assuming data are negative binomially distributed with mean *D*_*x*_. The reinfection hazard coefficient (*λ*) and the inverse of the negative binomial dispersion parameter (*κ*) were fitted to the data using a Metropolis-Hastings Monte Carlo Markov Chain (MCMC) estimation procedure implemented in the R Statistical Programming Language. We ran 4 MCMC chains with random starting values for a total of 10,000 iterations per chain, discarding the first 1,000 iterations (burn-in). Convergence was assessed using the Gelman-Rubin diagnostic (*26*).

#### Model-based projection

We used 1,500 samples from the joint posterior distribution of fitted model parameters to simulate possible reinfection time series under the null model, generating 100 stochastic realizations per parameter set. We then calculated projection intervals as the middle 95% of daily reinfection numbers across these simulations.

We applied this approach at the national and provincial levels.

#### Approach 2: Estimation of time-varying infection and reinfection hazards

We estimated the time-varying empirical hazard of infection as the daily incidence per susceptible individual. This approach requires reconstruction of the number of susceptible individuals through time. We distinguish between three “susceptible” groups: naive individuals who have not yet been infected (*S*_1_), previously infected individuals who had undetected infections at least 90 days ago and have not yet had a second infection 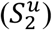, and previously infected individuals who had a prior positive test at least 90 days ago and have not yet had a second infection (*S*_2_). We estimate the numbers of individuals in each of these categories on day *t* as follows:

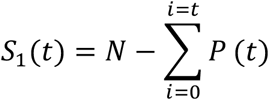

where *N* is the total population size and *P*(*t*) = *P*_*obs*_(*t*)/*p*_*obs*_ is the total number of primary infections on day *t*, of which *P*_*obs*_(*t*) were observed and *P*_*missed*_(*t*) = *P*(*t*) − *P*_*obs*_(*t*) were missed.

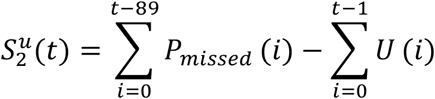

where 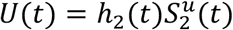 is the number of new infections among individuals whose first infection was missed. These individuals are assumed to experience the same infection hazard as individuals whose primary infection was diagnosed and who have not yet been reinfected, estimated as 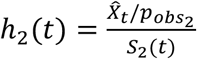. Because individuals are not eligible for reinfection until at least 90 days after their primary infection, we set *U*(*t*) = *h*_2_(*t*) = 0 when *t* < 90.

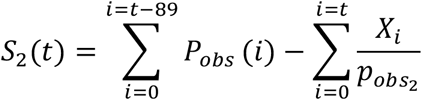

where 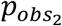 is the probability of detection for individuals who have had a previously identified infection, and *X*_*i*_ is the number of individuals with a second detected infection on day *i*. Only the possibility of second infections are accounted for in the model, which was developed to monitor reinfection risk against a background in which reinfections were rare.

This setup allows recursive calculation of *U*(*t*) and therefore 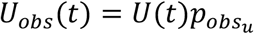, where 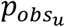 is the probability of a second infection being observed in an individual whose first infection was missed, and *P*_*obs*_(*t*) = *C*_*t*_ − *U*_*obs*_(*t*), where *C*_*t*_ is the number of individuals with their first positive test on day *t* (i.e., detected cases).

Individuals in 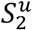 and *S*_2_ are assumed to experience the same daily hazard of reinfection, estimated as 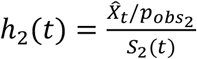. The daily hazard of infection for previously uninfected individuals is then estimated as 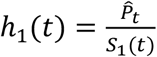.

If we assume that the hazard of infection is proportional to the 7-day moving average of infection incidence 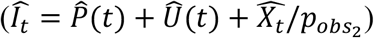, we can then examine the infectiousness of the virus through time as 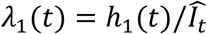 and 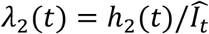. We constructed uncertainty intervals around *λ*_1_(*t*), *λ*_2_(*t*), and their ratio, taking into account both measurement noise and uncertainty in the observation parameters (see Supplementary Materials for details).

We also used this approach to construct a data set with the daily numbers of individuals eligible to have a primary infection (*S*_1_(*t*)) or suspected second infection (*S*_2_(*t*)) by wave. Wave periods were defined as the time surrounding the wave peak for which the 7-day moving average of case numbers was above 15% of the wave peak. We then analyzed these data using a generalized linear mixed model to estimate the relative hazard of infection in the population eligible for suspected second infection, compared to the hazard in the population not eligible for suspected second infection. For this analysis, we assume *p*_*obs*_ = 0.1 and 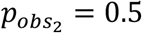, which falls within the plausible range of observation probabilities (see Figure S8).

Our primary regression model was a Poisson model with a log link function, groupinc = Poisson(*μ*):

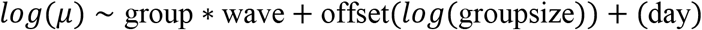

The outcome variable (groupinc) was the reconstructed daily number of observed infections in the two groups (*P*_*obs*_(*t*) and *X*_*t*_). Our main interest for this analysis was in whether the relative hazard was higher in the second wave, third wave, pre-wave period in which Omicron emerged, and/or fourth wave, relative to during the first wave, thus potentially indicating immune evasion. This effect is measured by the interaction term between group and wave. The offset term is used to ensure that the estimated coefficients can be appropriately interpreted as *per capita* rates. We used day as a proxy for force of infection and reporting patterns and examined models where day was represented as a random effect (to reflect that observed days can be thought of as samples from a theoretical population) and as a fixed effect (to better match the Poisson assumptions). As focal estimates from the two models were indistinguishable, we present only the results based on the random effect assumption.

## Data Availability

Data and code will be made available at https://github.com/jrcpulliam/reinfections. The following data are included in the repository:
- Counts of reinfections and primary infections by province, age group (5-year bands), and sex (M, F, U)
- Daily time series of primary infections and suspected reinfections by specimen receipt date (national)
- Model output: posterior samples from the MCMC fitting procedure and simulation results
Requests for additional data must be made in writing to the National Institute for Communicable Diseases, South Africa.

https://github.com/jrcpulliam/reinfections

## Acknowledgements

The authors wish to acknowledge the members of the NICD Epidemiology and Information Technology teams which curate, clean, and prepare the data utilized in this analysis.

## Epidemiology team

Andronica Moipone Shonhiwa, Genevie Ntshoe, Joy Ebonwu, Lactatia Motsuku, Liliwe Shuping, Mazvita Muchengeti, Jackie Kleynhans, Gillian Hunt, Victor Odhiambo Olago, Husna Ismail, Nevashan Govender, Ann Mathews, Vivien Essel, Veerle Msimang, Tendesayi Kufa-Chakezha, Nkengafac Villyen Motaze, Natalie Mayet, Tebogo Mmaborwa Matjokotja, Mzimasi Neti, Tracy Arendse, Teresa Lamola, Itumeleng Matiea, Darren Muganhiri, Babongile Ndlovu, Khuliso Ravhuhali, Emelda Ramutshila, Salaminah Mhlanga, Akhona Mzoneli, Nimesh Naran, Trisha Whitbread, Mpho Moeti, Chidozie Iwu, Eva Mathatha, Fhatuwani Gavhi, Masingita Makamu, Matimba Makhubele, Simbulele Mdleleni, Bracha Chiger, Jackie Kleynhans

## Information Technology team

Tsumbedzo Mukange, Trevor Bell, Lincoln Darwin, Fazil McKenna, Ndivhuwo Munava, Muzammil Raza Bano, Themba Ngobeni

We also thank Carl A.B. Pearson, Shade Horn, Youngji Jo, Belinda Lombard, Liz S. Villabona-Arenas, and colleagues in the South African COVID-19 Modelling Consortium and the SARS-CoV-2 variants research consortium in South Africa for helpful discussions during the development of this work. In addition, we acknowledge the Network for Genomic Surveillance - South Africa (NGS-SA) led by Prof Tulio de Oliveira for its role in discovery of the Omicron variant.

## Funding

JRCP and CvS are supported by the South African Department of Science and Innovation and the National Research Foundation. Any opinion, finding, and conclusion or recommendation expressed in this material is that of the authors and the NRF does not accept any liability in this regard. This work was also supported by the Wellcome Trust (grant number 221003/Z/20/Z) in collaboration with the Foreign, Commonwealth and Development Office, United Kingdom.

## Author contributions

Conceptualization - JP, CvS, JD, HM

Data collection, management, and validation - NG, KM, AvG, CC

Data analysis - JP, CvS, JD

Interpretation - JP, CvS, AvG, CC, MJG, JD, HM

Drafting the manuscript - JP, CvS, HM

Manuscript review, revision, and approval - all authors

Guarantor: HM

## Competing interests

All authors have completed the ICMJE uniform disclosure form. CC and AvG have received funding from Sanofi Pasteur in the past 36 months. JRCP and KM serve on the Ministerial Advisory Committee on COVID-19 of the South African National Department of Health. The authors have declared no other relationships or activities that could appear to have influenced the submitted work.

## Ethics statement

Ethical approval: This study has received ethical clearance from University of the Witwatersrand (Clearance certificate number M210752, formerly M160667) and approval under reciprocal review from Stellenbosch University (Project ID 19330, Ethics Reference Number N20/11/074_RECIP_WITS_M160667_COVID-19).

## Data availability

Data and code will be made available at https://github.com/jrcpulliam/reinfections as release 3. The following data will be included in the repository:

- Counts of reinfections and primary infections by province, age group (5-year bands), and sex (M, F, U)
- Daily time series of primary infections and suspected reinfections by specimen receipt date (national)

All other data are covered by a non-disclosure agreement and cannot be released by the authors. Requests for additional data must be made in writing to the National Institute for Communicable Diseases, South Africa. Model output (posterior samples from the MCMC fitting procedure and simulation results) will be made available via Zenodo due to large file sizes.

This work is licensed under a Creative Commons Attribution 4.0 International (CC BY 4.0) license, which permits unrestricted use, distribution, and reproduction in any medium, provided the original work is properly cited. To view a copy of this license, visit https://creativecommons.org/licenses/by/4.0/. This license does not apply to figures/photos/artwork or other content included in the article that is credited to a third party; obtain authorization from the rights holder before using such material.

## Supplementary Materials

### Supplementary Text

#### 1. Supplementary Methods

##### 1.1 Data validation and known data issues

To assess validity of the data linkage procedure and thus verify whether individuals identified as having suspected reinfections did in fact have positive test results at least 90 days apart, we conducted a manual review of a random sample of suspected second infections occurring on or before 20 January 2021 (n=585 of 6026; 9.7%). This review compared fields not used for linkages (address, cell-phone numbers, email addresses, facility, and health-care providers) between records in the NMC-SS and positive test line lists. Where uncertainty remained and contact details were available, patients or next-of-kin were contacted telephonically to verify whether the individual had received multiple positive test results.

Of the 585 randomly selected individuals with possible reinfections in the validation sample, 562 (96%) were verified as the same individual based on fields not used to create the linkages; the remaining 23 (4%) were either judged not a match or to have insufficient evidence (details captured by the clinician or testing laboratory) to determine whether the records belonged to the same individual.

Between 5 and 11 December 2022, server issues at the National Health Laboratory Service’s Central Data Warehouse prevented one of the identifiers used in the probabilistic linkages from being pulled through into the dataset. Retrospective evaluation of the use of the identifier revealed that it had been required for 0.3% of the links made prior to this time. The impact of this data discrepancy is therefore thought to be minimal.

##### 1.2 Descriptive analysis

We compared the age, gender, and province of individuals with suspected reinfections to individuals eligible for reinfection (i.e., who had a positive test result at least 90 days prior to 31 January 2022).

We did not calculate overall incidence rates by wave because the force of infection is highly variable in space and time, and the period incidence rate is also influenced by the temporal pattern of when people become eligible for reinfection. Incidence rate estimates would therefore be strongly dependent on the time frame of the analysis and not comparable to studies from other locations or time periods.

##### 1.3 Construction of uncertainty intervals for hazard coefficients and hazard ratio

The uncertainty intervals shown in Figure 5 were constructed to take into account both measurement error and uncertainty in the assumed observation probabilities. To capture uncertainty in the observation probabilities, we uniformly sampled 1,000 values from the polygon of plausible values for *p*_*obs*_ and 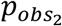 (Figure S8). For each parameter combination, we used the model described in the main text (approach 2) to reconstruct the numbers of primary and second infections by day, as well as the relevant susceptible populations at risk. We then used each reconstructed data set to construct a 95% confidence interval for the associated Poisson rate, after Sahai and Kurshid (*1*), and for the associated incidence rate ratio, after Ulm (*2*). The confidence limits for the hazard coefficients were approximated by dividing the confidence limits for the Poisson rates by the reconstructed value of the total incidence for each reconstructed data set.

The final uncertainty intervals were then constructed from the distribution of confidence limits based on the 1,000 reconstructed data sets. The median value presented in Figure 5 is the median estimate from across the data sets, and the confidence limits represent the 2.5% and 97.5% quantiles of the lower and upper confidence limits, respectively.

#### 2. Supplementary Results

##### 2.1 Distribution of suspected reinfections by province

Suspected reinfections were identified in all nine provinces (Figure S1). The reinfection rate was highest in Western Cape, where 20,952 of 516,857 eligible primary infections (4.05%) had suspected reinfections and lowest in Northern Cape (2,464 of 92,718; 2.66%). For comparison, the national reinfection rate was 92,718; 3.58% (105,323 of 2,942,248 eligible primary infections). Numbers for all provinces are provided in Table S1.

**Fig. S1.**
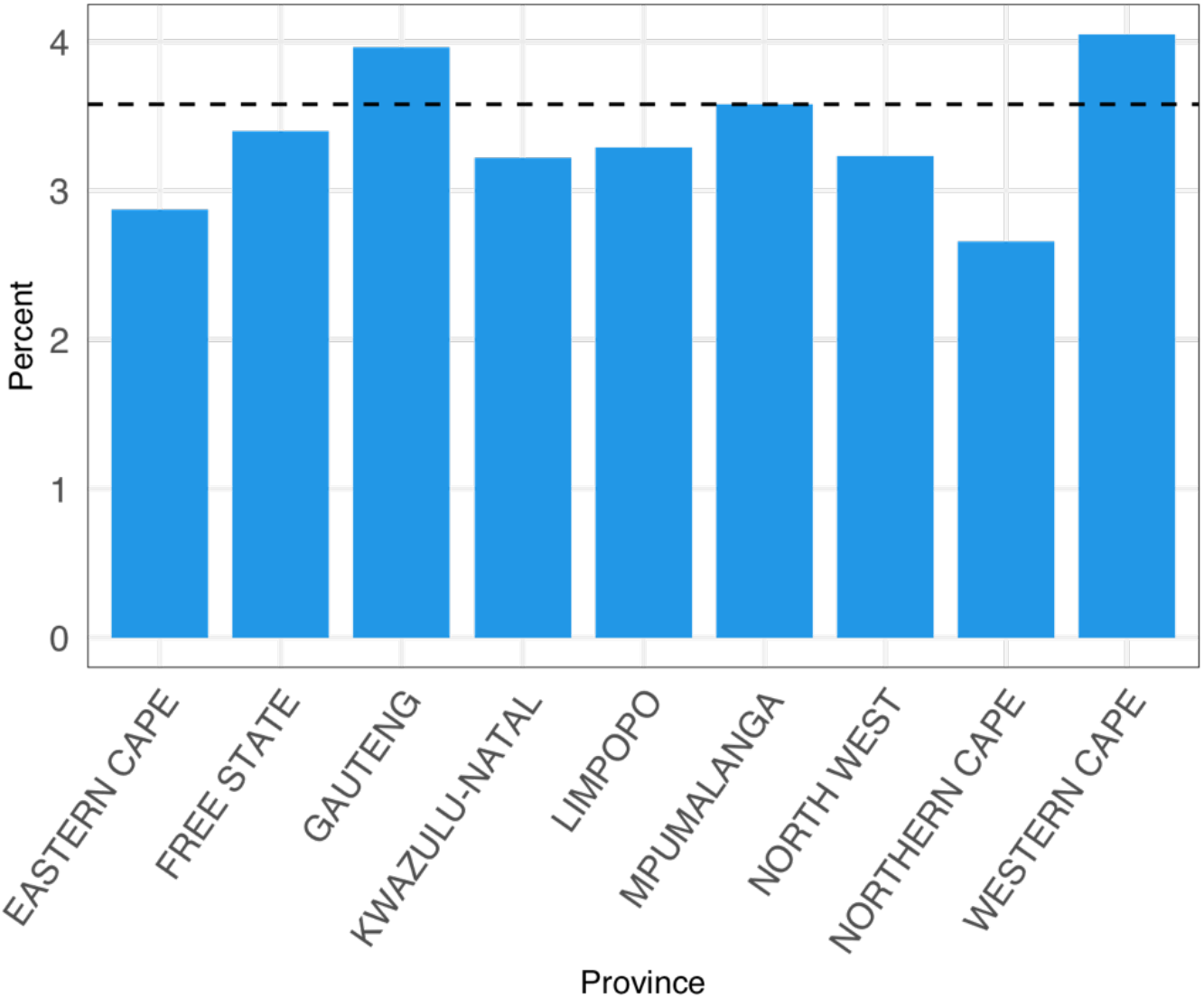
Descriptive analysis of suspected reinfections: Percentage of eligible primary infections with suspected reinfections, by province.

**Table S1.** Distribution of suspected reinfections by province, South Africa, March 2020 to January 2022.

| Province | No reinfection | One reinfection | Two reinfections | Total |
| --- | --- | --- | --- | --- |
| EASTERN CAPE | 284,397 | 8,285 | 127 | 292,809 |
| FREE STATE | 159,055 | 5,505 | 87 | 164,647 |
| GAUTENG | 891,193 | 36,149 | 597 | 927,939 |
| KWAZULU-NATAL | 501,040 | 16,409 | 256 | 517,705 |
| LIMPOPO | 119,323 | 3,968 | 87 | 123,378 |
| MPUMALANGA | 147,573 | 5,394 | 92 | 153,059 |
| NORTH WEST | 148,183 | 4,845 | 106 | 153,134 |
| NORTHERN CAPE | 90,254 | 2,438 | 26 | 92,718 |
| WESTERN CAPE | 495,905 | 20,552 | 400 | 516,857 |
| UNKNOWN | 2 | 0 | 0 | 2 |
| **Total** | **2,836,925** | **103,545** | **1,778** | **2,942,248** |

##### 2.2 Breakdown of suspected reinfections by sex and age group

Among 2,878,217 eligible primary infections with both age and sex recorded, 62,690 of 1,630,428 females (3.85%) and 42,099 of 1,247,789 males (3.37%) had suspected reinfections. Relative to individuals with no identified reinfection, reinfections were concentrated in adults between the ages of 20 and 55 years (Figure S2). Numbers for all age group-sex combinations are provided in Table S2.

**Fig. S2.**
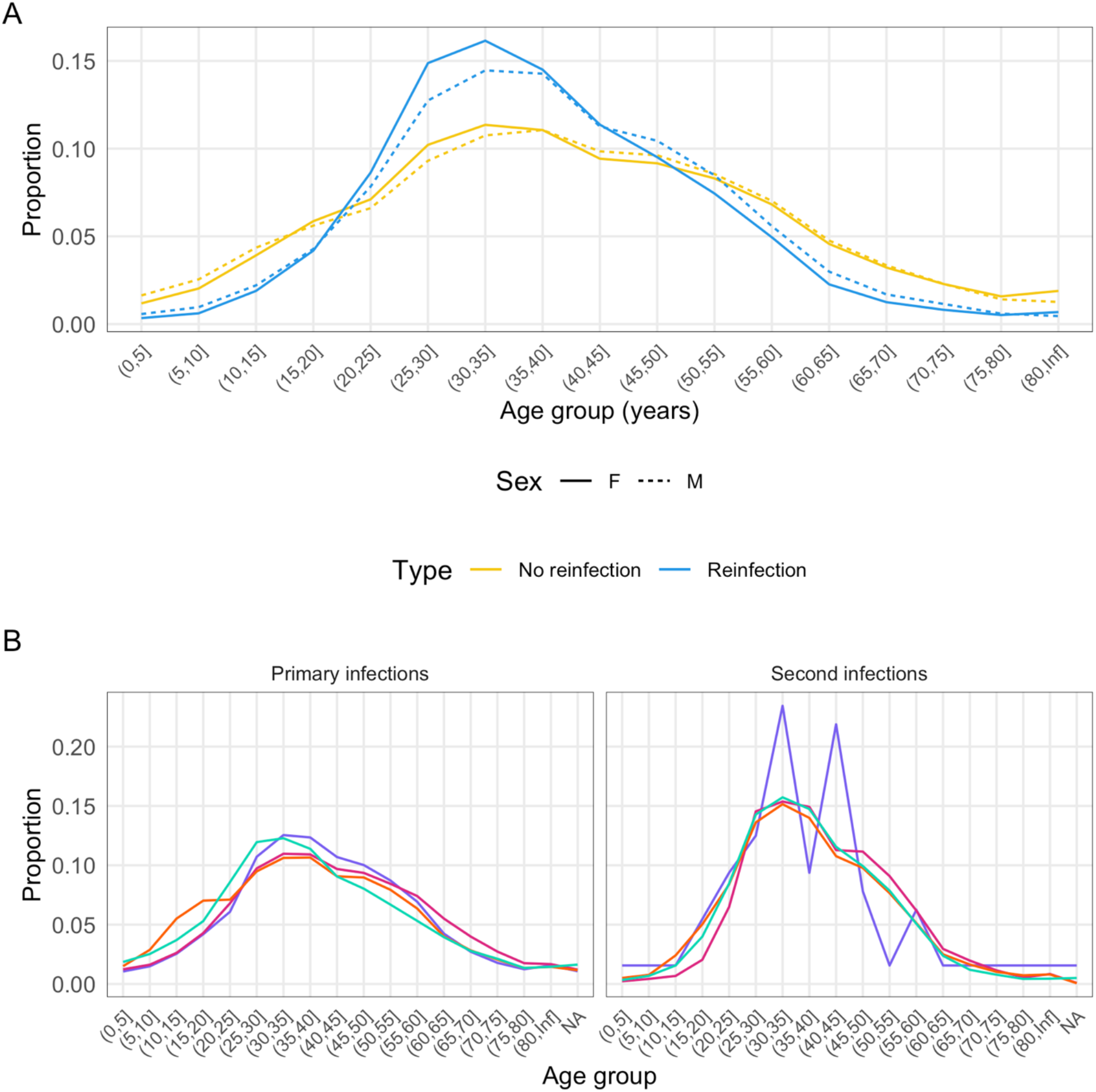
Descriptive analysis of suspected reinfections: A: Age distribution of individuals with suspected reinfections (blue) versus eligible individuals with no detected reinfection (yellow), by sex. Solid lines indicate females; dashed lines indicate males. B: Age distribution of primary infections (left) and second infections (right) by wave (purple = wave 1, pink = wave 2, orange = wave 3, turquoise = wave 4).

**Table S2.** Breakdown of suspected reinfections by sex and age group (years), South Africa, March 2020 to January 2022.

| **Sex** | **Age group** | **No reinfection** | **One reinfection** | **Two reinfections** | **Total** |
| --- | --- | --- | --- | --- | --- |
| F | (0,20] | 203,687 | 4,361 | 50 | 208,098 |
| F | (20,40] | 623,098 | 33,299 | 656 | 657,053 |
| F | (40,60] | 528,525 | 20,505 | 353 | 549,383 |
| F | (60,80] | 182,785 | 3,004 | 33 | 185,822 |
| F | (80,Inf] | 29,643 | 426 | 3 | 30,072 |
| M | (0,20] | 170,647 | 3,353 | 27 | 174,027 |
| M | (20,40] | 454,922 | 20,364 | 397 | 475,683 |
| M | (40,60] | 422,514 | 14,841 | 220 | 437,575 |
| M | (60,80] | 142,421 | 2,673 | 31 | 145,125 |
| M | (80,Inf] | 15,186 | 188 | 5 | 15,379 |
| **Total** |  | **2,773,428** | **103,014** | **1,775** | **2,878,217** |

**Fig. S3.**
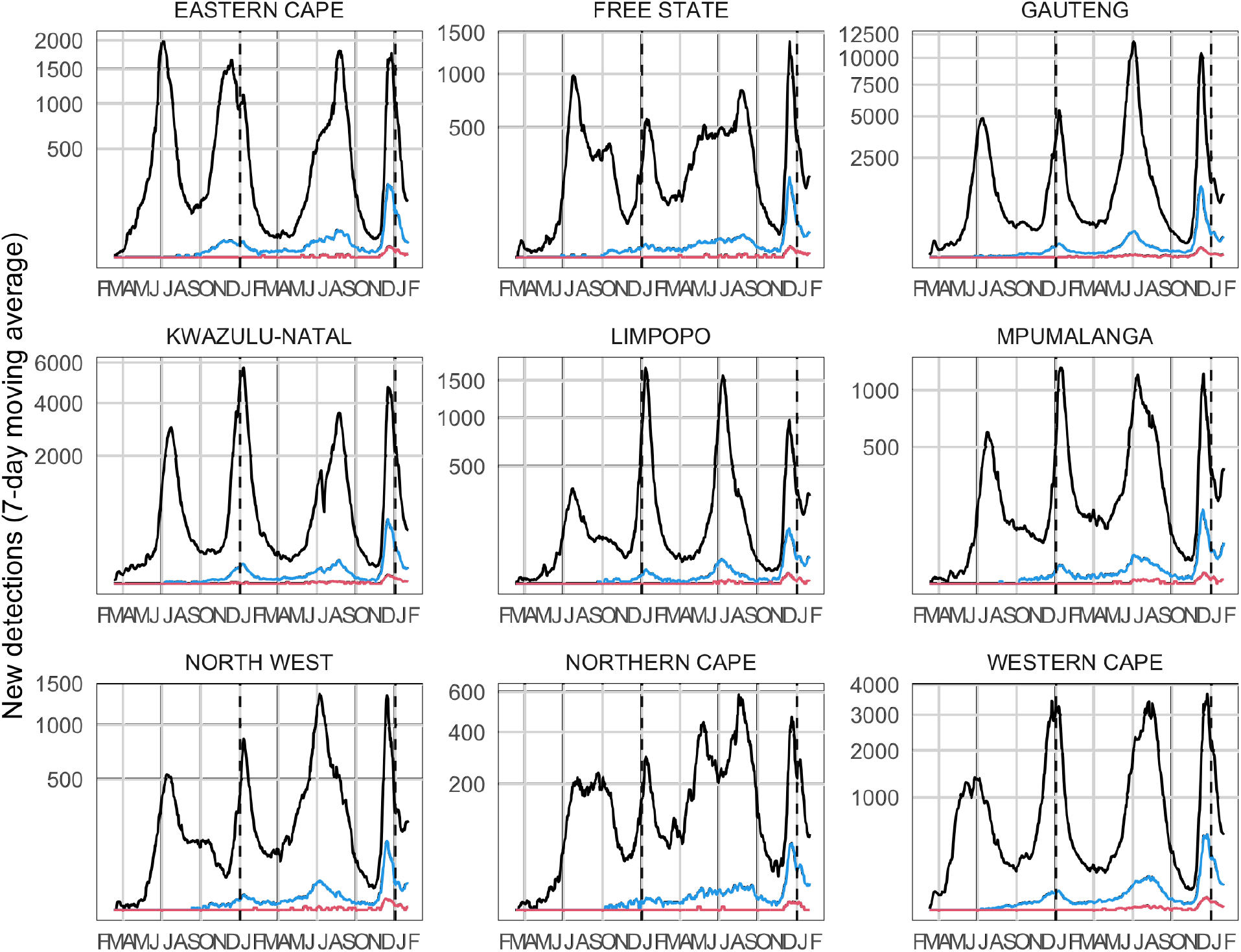
Number of detected primary infections (black), suspected reinfections (blue), and suspected third infections (red), by province. Lines represent 7-day moving averages. The y-axes are shown on a square root scale.

**Fig. S4.**
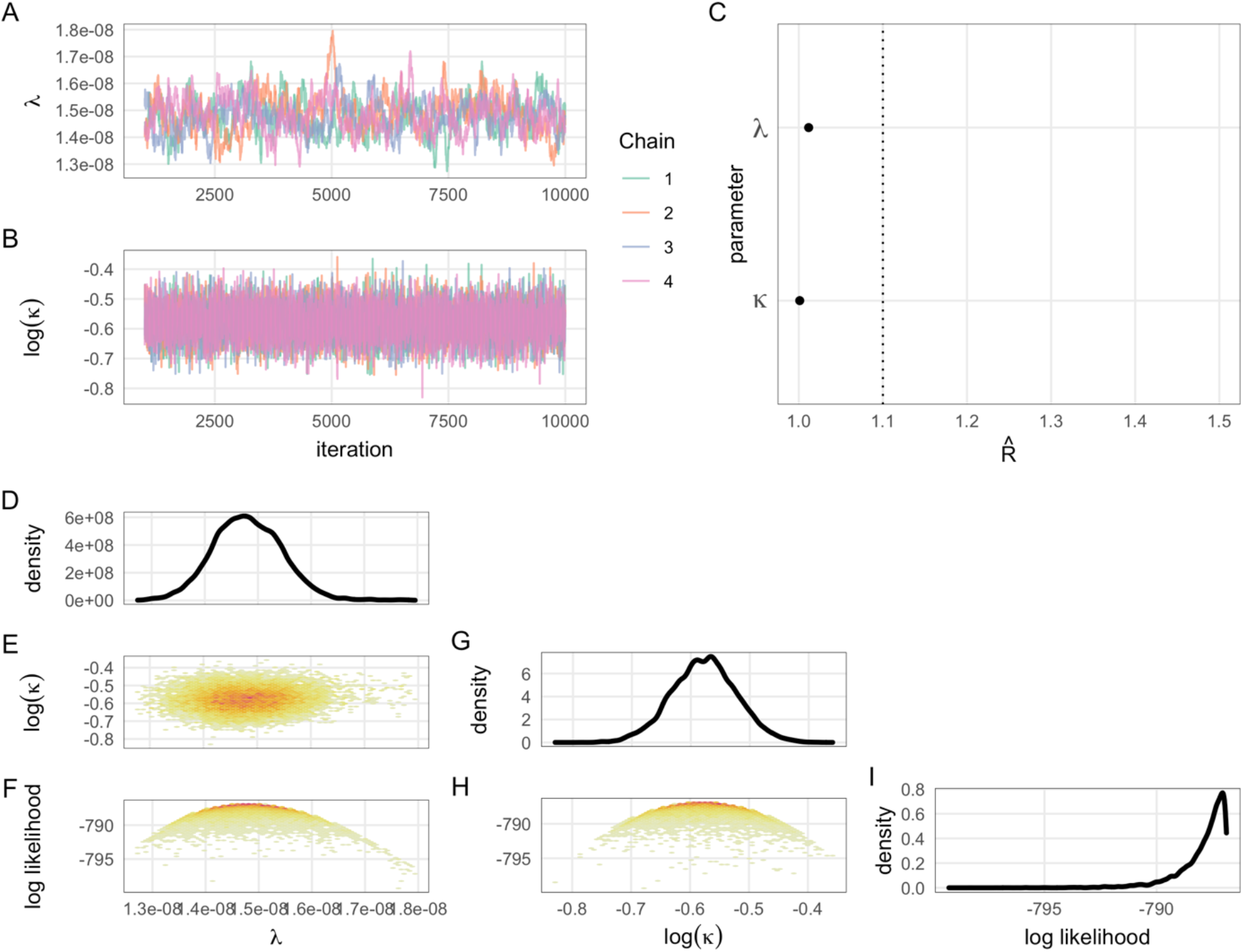
Convergence diagnostics and density of the posterior distribution for MCMC fits (approach 1). A and B: MCMC chains for each parameter. C: Gelman-Rubin values (a.k.a. potential scale reduction factors) for each parameter; values less than 1.1 indicate sufficient mixing of chains to suggest convergence. D, G, I: posterior density for each parameter and the log likelihood. E, F, H: 2-D density plots showing correlations between parameters and the log likelihood.

**Fig. S5.**
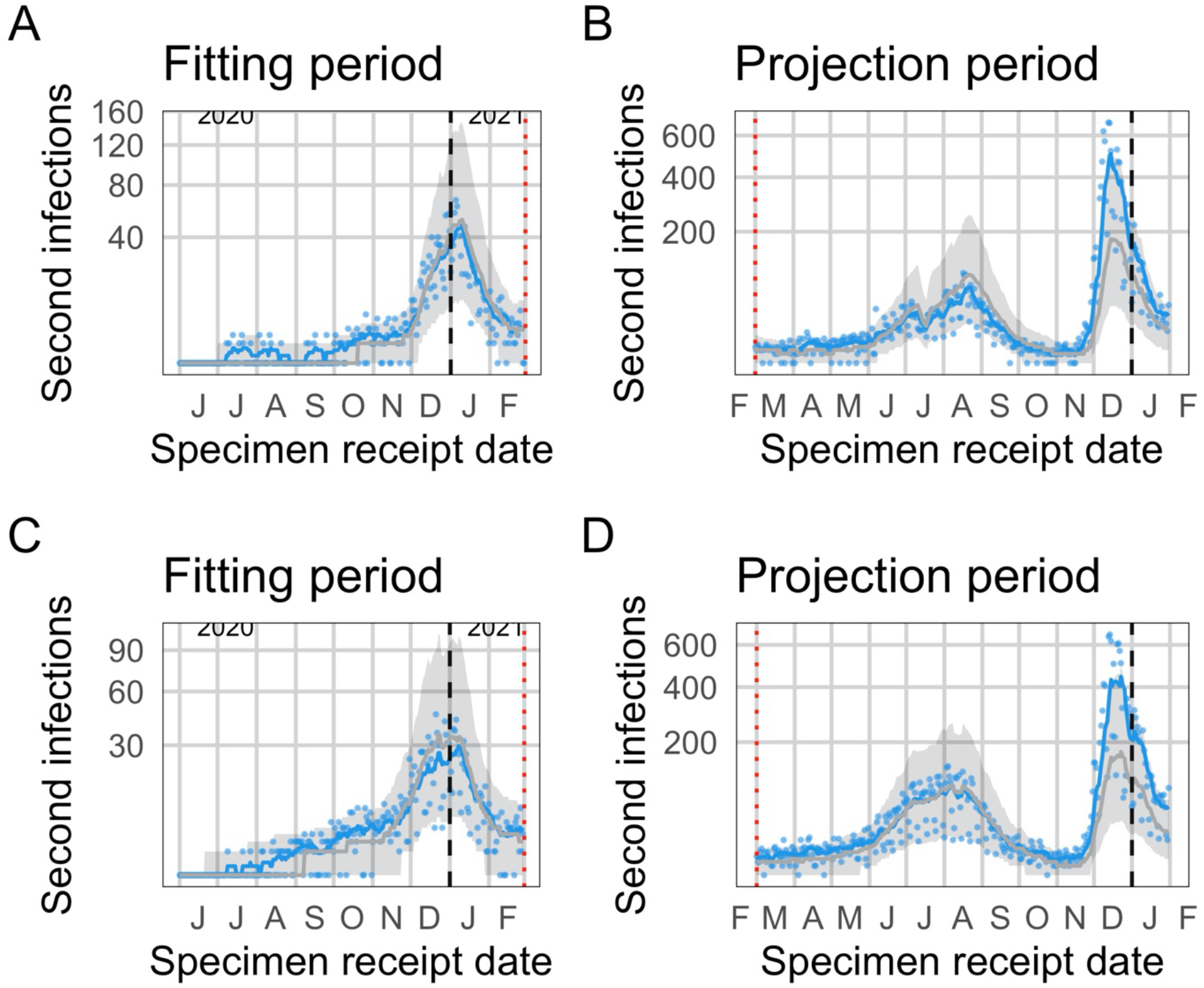
Observed and expected temporal trends in reinfection numbers, for the second and third most populous provinces. Blue lines (points) represent the 7-day moving average (daily values) of suspected reinfections. Grey lines (bands) represent mean predictions (95% projection intervals) from the null model. A and B: KwaZulu-Natal. C and D: Western Cape.

**Fig. S6.**
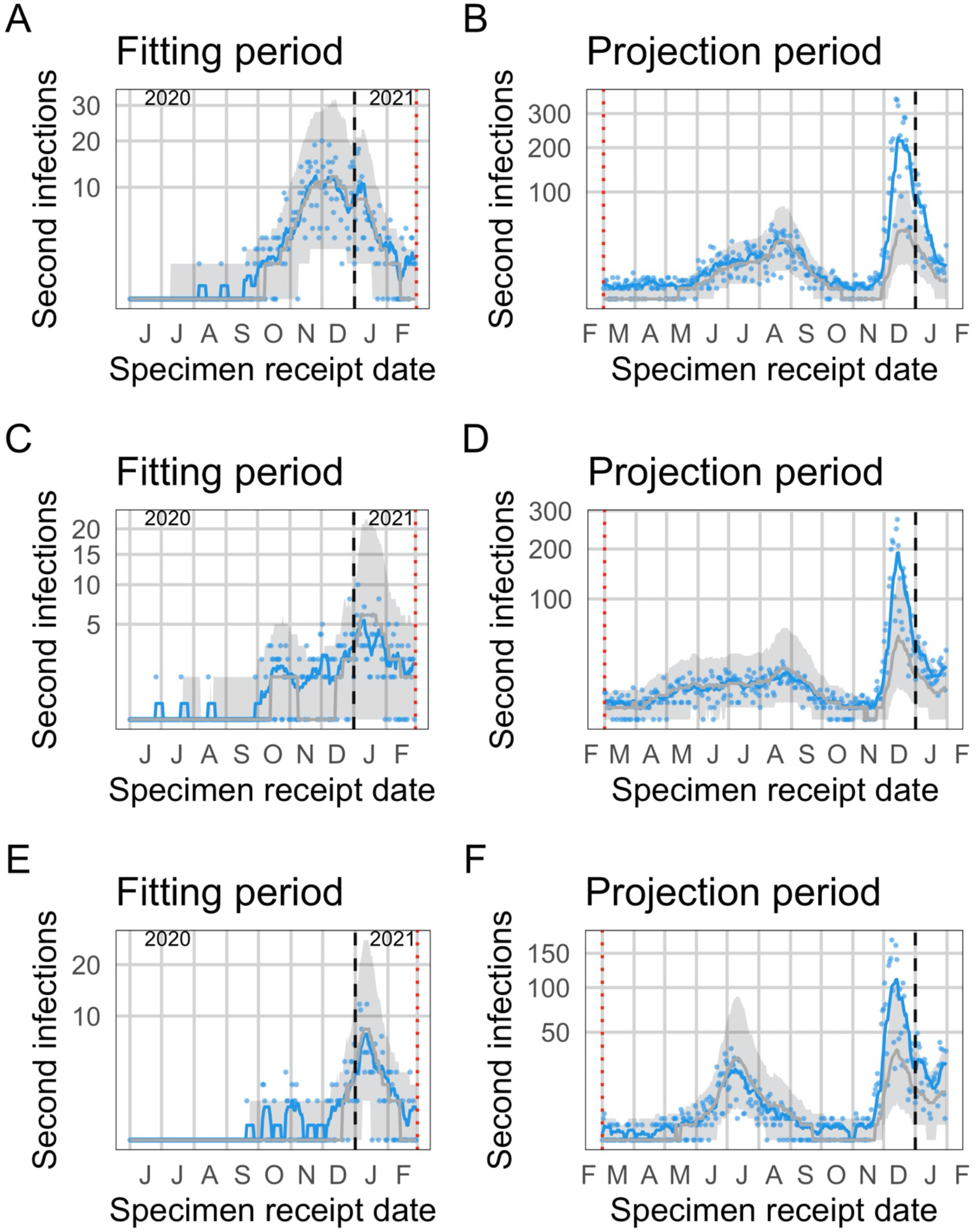
Observed and expected temporal trends in reinfection numbers. Blue lines (points) represent the 7-day moving average (daily values) of suspected reinfections. Grey lines (bands) represent mean predictions (95% projection intervals) from the null model. A and B: Eastern Cape. C and D: Free State, E and F: Limpopo.

**Fig. S7.**
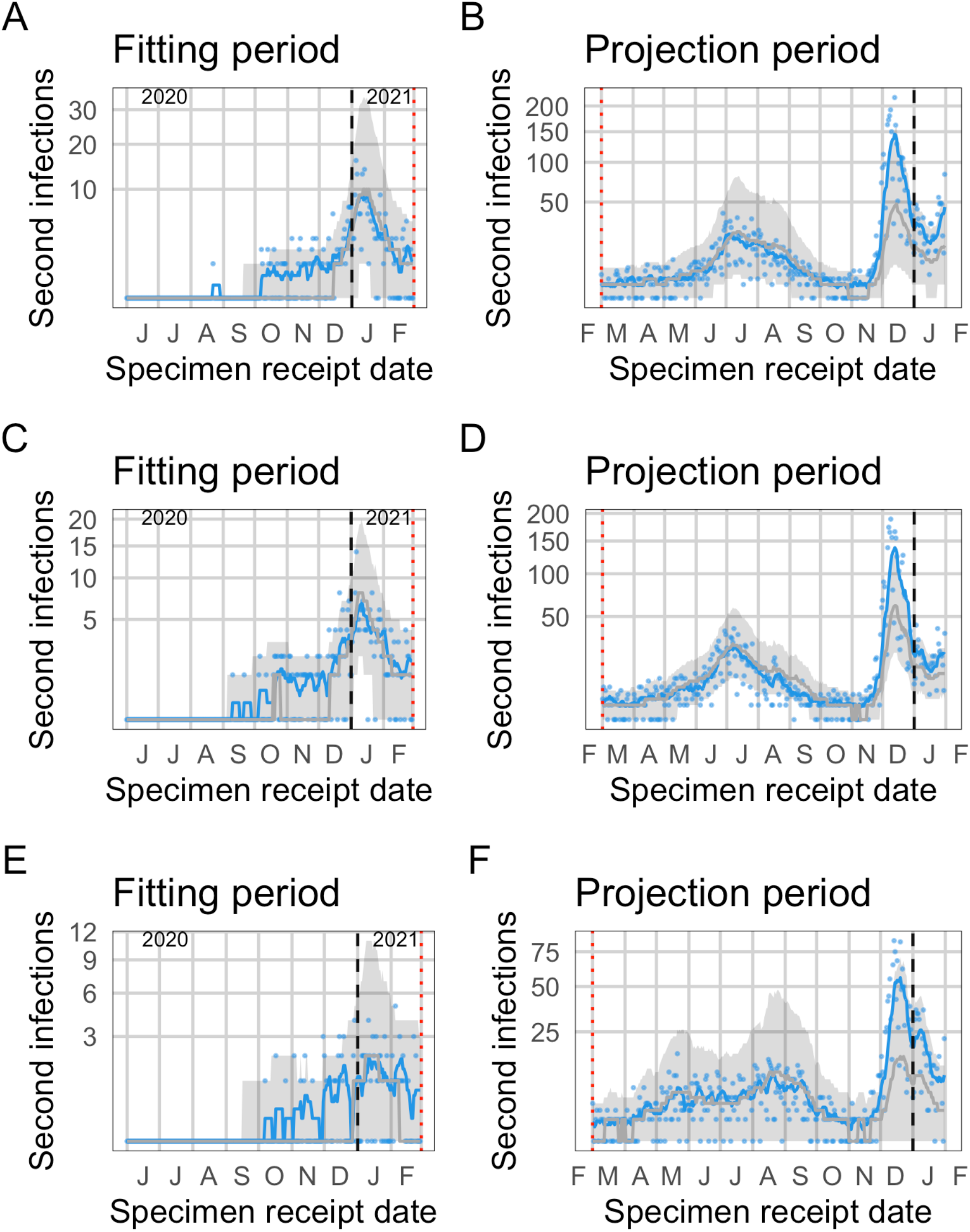
Observed and expected temporal trends in reinfection numbers. Blue lines (points) represent the 7-day moving average (daily values) of suspected reinfections. Grey lines (bands) represent mean predictions (95% projection intervals) from the null model. A and B: Mpumalanga. C and D: North West, E and F: Northern Cape.

**Fig. S8.**
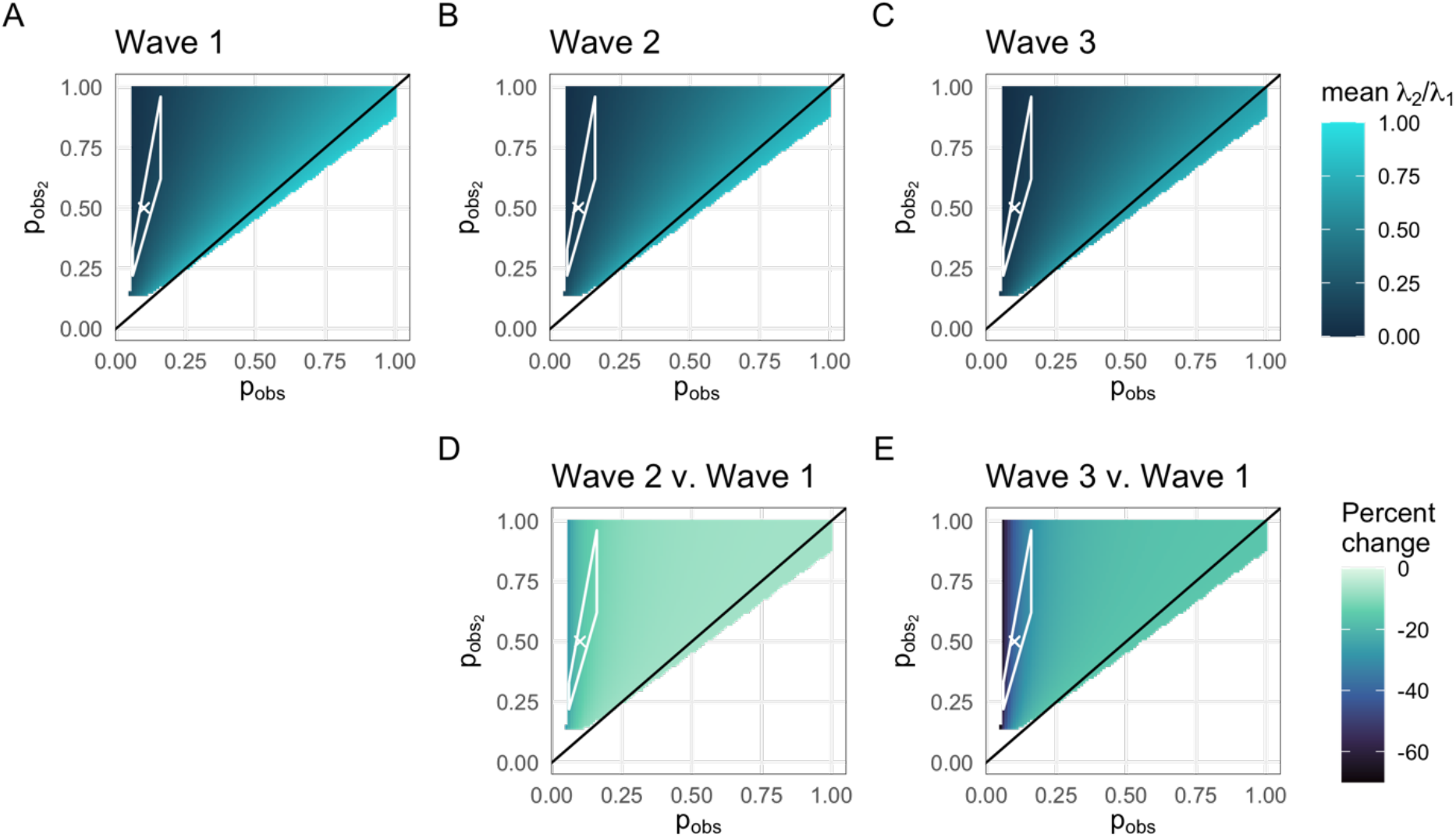
Sensitivity analysis of hazard ratio estimates to assumed observation probabilities for primary and second infections. Estimates are shown for the full range of probabilities for which the overall mean relative hazard is between 0 and 1. The white polygon encloses the most plausible estimates (i.e. consistent with relative reinfection risk observed in the SIREN study (*3*) and observation probabilities for primary infection consistent with estimates based on seroprevalence data (*4*)). For all parameter combinations in the plausible range, 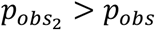, suggesting that having had a previous test is a good marker for who will test again. Top: Mean relative empirical hazard for reinfections versus primary infections in each wave, as a function of assumed observation probabilities for primary infections (*p*_*obs*_) and reinfections 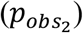. A: wave 1, B: wave 2, C: wave 3. Bottom: Percent change in the mean relative empirical hazard for reinfections versus primary infections in waves 2 (D) and 3 (E) relative to wave 1, as a function of assumed observation probabilities for primary infections (*p*_*obs*_) and reinfections 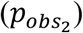.

**Fig. S9.**
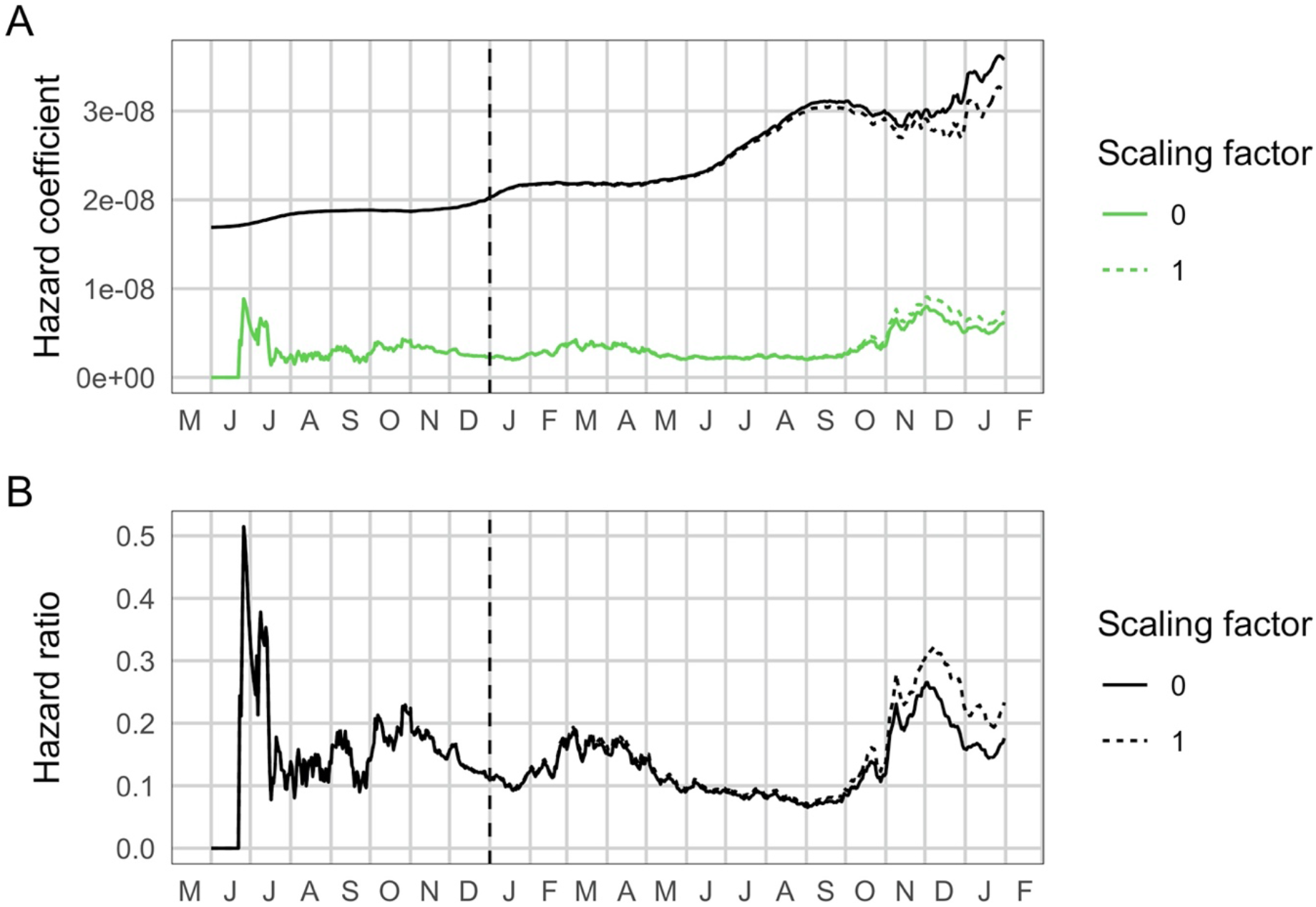
Sensitivity analysis of empirical hazard ratio estimates to assumed observation probability for second infections among individuals whose first infection was undetected 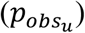. In the main analysis we assume that this observation probability is equivalent to other individuals who have not yet had a detected infection (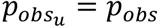, or a scaling factor of 1). Here, we compare this to the case when this probability is equal to 0 (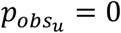, or a scaling factor of 0). We consider these to be bounding cases. The figure shown here is for *p*_*obs*_ = 0.1 and 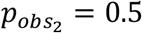.

## Notes

### Funding Statement

This work was supported by the South African Department of Science and Innovation and the National Research Foundation and the Wellcome Trust (grant number 221003/Z/20/Z) in collaboration with the Foreign, Commonwealth and Development Office, United Kingdom.

### Author Declarations

This study has received ethical clearance from University of the Witwatersrand (Clearance certificate number M210752, formerly M160667) and approval under reciprocal review from Stellenbosch University (Project ID 19330, Ethics Reference Number N20/11/074_RECIP_WITS_M160667_COVID-19).

### Summary of Updates

This version of the manuscript has been revised with the following major changes: (1) we now present data through 31 January 2022, covering the full Omicron wave in South Africa, and (2) we fixed an inconsistency in the calculation of the time-varying infection and reinfection hazard coefficients that enhanced the apparent decrease in primary infection hazard. The latter change has a quantitative impact on the results, but the conclusions remain unchanged.

